# EQUALITY OF OPPORTUNITY FOR TIMELY DEMENTIA DIAGNOSIS (EQUATED): A QUALITATIVE STUDY OF HOW PEOPLE FROM MINORITIZED ETHNIC GROUPS EXPERIENCE THE EARLY SYMPTOMS OF DEMENTIA AND SEEK HELP

**DOI:** 10.1101/2024.02.12.24302683

**Authors:** Christine Carter, Moïse Roche, Elenyd Whitfield, Jessica Budgett, Sarah Morgan-Trimmer, Sedigheh Zabihi, Yvonne Birks, Fiona Walter, Mark Wilberforce, Jessica Jiang, Refah Z Ahmed, Wesley Dowridge, Charles R Marshall, Claudia Cooper

## Abstract

**Introduction:** People from minoritized ethnic groups are diagnosed with dementia later in the disease. We explored pathways that may determine the timing of diagnoses in an ethnically diverse, urban area.

**Methods:** We conducted 61 semi-structured interviews: 10 community-dwelling older people from minoritized ethnic backgrounds with diagnosed and undiagnosed dementia (mean age = 72 years; males = 5/10), 30 family members (51, 10/30), 16 health or social care professionals (42; 3/15) three paid carers and two interpreters for people with dementia. We used reflexive thematic analysis, and the Model of Pathways to Treatment to consider diagnostic delay.

**Findings:** We identified three themes: (1) ***Cultural identity and practices shape responses***: gendered expectations that families relieve elders of household roles reduce awareness or concern when functioning declines; expectations that religious practices are maintained mean problems doing so triggers help-seeking. Second generation family members often held insider and outsider identities, balancing traditional and Western perspectives. (2) ***Becoming like a tourist:*** daily experiences became unfamiliar for people developing dementia in an adopted country, sometimes engendering a need to reconnect with a home country. For professionals and interpreters, translating meanings faithfully, and balancing relatives’ and clients’ voices, were challenging. (3) ***Naming and conceptualising dementia*:** the term dementia was stigmatised, with cultural nuances in how it was understood; initial presentations often included physical symptoms with cognitive concerns.

**Conclusion:** Greater understanding of dilemmas faced by minoritized ethnic communities, closer collaboration with interpreters and workforce diversity could reduce time from symptom appraisal to diagnosis, and support culturally competent diagnostic assessments.

## Introduction

People typically wait over three years from symptom onset to dementia diagnosis (1). Timely diagnosis empowers people to understand symptoms, make plans and access symptomatic treatments. People living in less affluent areas and from ethnic and other minoritized groups are typically diagnosed with dementia later and with processes characterised by less accuracy and precision (2–5). Qualitative studies in Norway (6,7) and Canada (8) have suggested possible reasons for this, including inappropriate attribution of symptoms to normal ageing, stigma, lack of professional knowledge regarding diagnostic tools and challenges from language barriers. In this context symptom attribution refers to the personal and social construction of meaning surrounding dementia, and is highly context dependent (29). Among Indian people with mild dementia and family carers in New Zealand, dementia was often attributed to “karma”, engendering feelings of acceptance, and delaying help seeking (9). McCleary et al (8) described how South Asian family carers often modified environments to account for symptoms rather than seek medical help. Sagbakken describes the challenge to services “to become familiar with each person’s way of being ill, on a cultural and individual level”(10).

To our knowledge, only one UK published study has qualitatively interviewed people from ethnic minoritized ethnic groups affected by dementia about reasons for diagnostic delay. Family carers from South Asian communities described how early cognitive changes were often initially conceptualised as physical or affective changes (14).

We used the Model of Pathways to Treatment (15) (Table 1) as a theoretical framework, to consider the pathways which determine the timing of dementia diagnosis in an ethnically diverse, London area. This model addresses the complexity and dynamic pathways which shape diagnosis and treatment delay (15). We included people from minoritized ethnic communities living in East London, where the most common minority ethnic groups are Asian or Asian British, and Black or Black British, Caribbean or African. While experiences may differ between minority ethnic cultures, there is a common risk of being diagnosed with dementia later. We sought to explore how experiences of immigration and the effects of globalization on family life and traditional kinships might explain delayed diagnosis and engagement with and by health services.

**Table 1:** Sociodemographic characteristics of participants.

| Demographic |  | People with dementia (n=10) | Family carers (n=30) | Paid carers (n=3) | Health care Professionals (n=16) | Interpreters (n=2) |
| --- | --- | --- | --- | --- | --- | --- |
| Ethnicity: Asian | Other |  | 2 |  |  |  |
|  | Bangladeshi | 4 | 13 | - | 1 |  |
|  | British | - |  | - | 2 | 1 |
|  | Chinese | 1 | 2 | - | 1 |  |
|  | Pakistani | - | 1 | - | - |  |
|  | Philippines | - |  | - | 1 |  |
|  | Vietnamese | - | 1 | - | 1 |  |
| Black | African | 2 | - | - | 2 | 1 |
|  | British | - | 1 | - | - |  |
|  | Caribbean | 2 | 5 | 1 | - |  |
| Mixed/ Other | Bangladeshi/British | - | 2 | - | - |  |
|  | Bengali/Italian | - | 2 | - | - |  |
|  | Black British (mixed) | - | - | - | 3 |  |
|  | Black Caribbean/White | 1 | 1 | - | - |  |
|  | Caribbean/Vietnamese | - | - | 1 | - |  |
|  | Somalian | - |  | 1 |  |  |
| White | British | - | - | - | 3 |  |
|  | European<sup>5</sup> | - | - | - | 2 |  |
| Sex | Female | 5 | 20 | 3 | 15 | 2 |
|  | Male | 5 | 10 | - | 3 | 0 |
| Age | Mean (SD) in years | 72 (14) | 51 (17) | 50 (15) | 42 (9) | 39(3) |
| Marital status | Married/civil partnership | 4 | 17 | - | - |  |
|  | Single | 3 | 11 | - | - |  |
|  | Widowed | 2 | 1 | - | - |  |
|  | Divorced | 1 | 1 | - | - |  |
| Living with | Alone | 3 | 5 | - | - |  |
|  | Partner/spouse | 3 | 6 | - | - |  |
|  | Parent(s)<sup>6</sup> | - | 13 | - | - |  |
|  | Children | 3 | 6 | - | - |  |
|  | Daughter-in-law | 1 |  | - | - |  |
| Accommodation | Council rented | - | 4 | 2 | - |  |
|  | Housing association | 4 | 10 | 1 | - |  |
|  | Private rented | - | 8 | - | - |  |
|  | Owner-occupied | 5 | 8 | - | - |  |
|  | Assisted living | 1 | - | - | - |  |
| Education | Degree/ postgraduate | 2 | 14 | - | - |  |
|  | Aged 16/18+ qualifications | 2 | 12 | 3 | - |  |
|  | No formal qualifications | 6 | 4 | - | - |  |
| Professional Roles | Nurse | - | - | - | 5 |  |
|  | General Practitioner | - | - | - | 3 |  |
|  | Manager | - | - | - | 1 |  |
|  | Occupational Therapist | - | - | - | 1 |  |
|  | Psychiatrist | - | - | - | 1 |  |
|  | Psychologist | - | - | - | 1 |  |
|  | Geriatrician | - | - | - | 1 |  |
|  | Therapist | - | - | - | 2 |  |
|  | Neurologist | - | - | - | 1 |  |
| Dementia subtype | Alzheimer's Disease | 1 | 10 | - | - |  |
|  | Vascular dementia | - | 5 | - | - |  |
|  | Mixed dementia | - | 3 | - | - |  |
|  | Memory problems<sup>8</sup> | 5 | 7 | - | - |  |
|  | Not known<sup>9</sup> | 4 | 5 | - | - |  |
| First language | English | 5 | 14 | 1 | 10 |  |
|  | Bengali | 4 | 12 | - | 1 |  |
|  | Cantonese | 1 | 2 | - | 0 | 1 |
|  | French | - | - | - | 1 |  |
|  | Polish | - | - | - | 1 |  |
|  | Swedish | - | - | - | 1 |  |
|  | Vietnamese | - | 1 | 1 | 0 | 1 |
|  | Somalian | - | - | 1 | - |  |
|  | Tangelo | - | - | - | 1 |  |
|  | Urdu | - | 1 | - | - |  |
|  | Zulu | - | - | - | 1 |  |

**Table 2:**
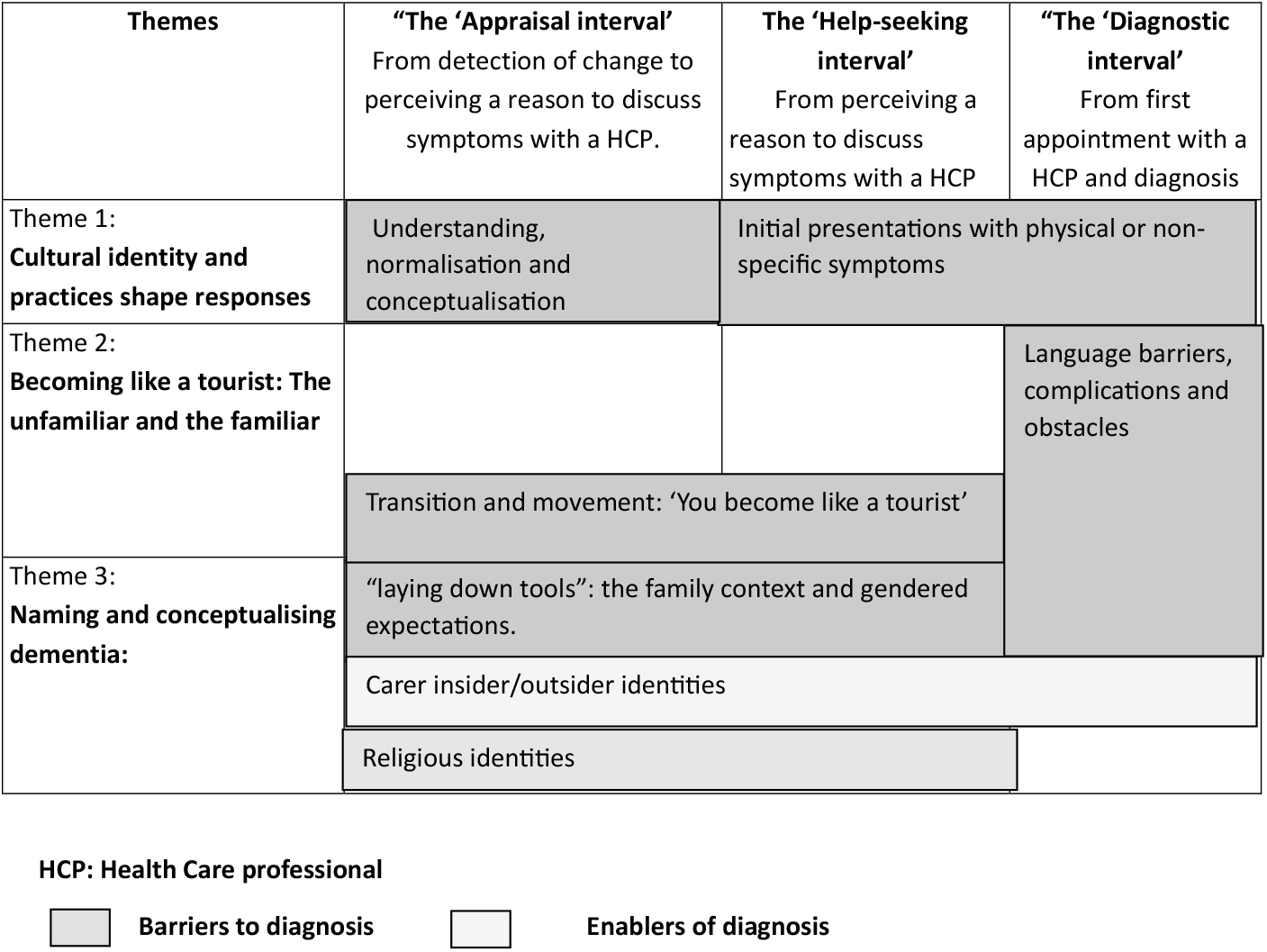
How themes fit in the Model of Pathways to Treatment (15) framework.

We aimed to explore the complex, dynamic pathways that determine the timing of diagnoses in an urban area of high ethnic density. Our secondary aim was to understand how services should be adapted to account for different styles of appraisal of dementia symptoms and help-seeking and be more culturally competent.

## Method

### Study design

Camden & King’s Cross Research Ethics Committee (REC: 23/YH/0034) approved this qualitative interview study on 1.3.23.

### Study setting and population

We planned our sample size to ensure sufficient diversity (16); with the *a priori* intention to include 40-60 people. We recruited participants from the non-statuary services and National Health Service (NHS) primary and secondary care organisations. We used a sampling frame to purposively recruit older people (aged 60+) living in East London who were experiencing symptoms associated with dementia, for diversities of culture, gender, ethnicity, disability, and other experiences (migration, deprivation). We included participants with a dementia diagnosis or symptoms of dementia (Noticeable Problems Checklist score of 5+ (17)) without a formal diagnosis. We also sought to recruit their family members. We recruited a range of professionals who diagnosed dementia or supported people to access diagnostic services in East London, including professional interpreters and paid care workers. Health care professionals were recruited for role diversity across primary and secondary NHS care and non-statutory organisations.

### Procedures

CCa, MR and EW (experienced qualitative researchers) conducted semi-structured interviews between April - September 2023 via video-call or face-to-face, as interviewees preferred. All interviews were recorded and professionally transcribed. People with dementia were interviewed with carers if they preferred this.

We consulted patient and public collaborators, and the clinical and academic team to develop topic guides based around our aim (above) and accounting for our theoretical framework (15) (Appendix 1). They explored experiences of prodromal dementia; initial symptoms, patterns of help-seeking and reasons underpinning this, and among those in receipt of a dementia diagnosis, pathways to diagnosis.

We only interviewed participants who could give informed consent. We asked people living with dementia to consent to interviews with family and paid carers about their symptoms and care. For those who did not have capacity, we identified a personal consultee, who we asked to complete a consultee declaration form, in line with the Mental Capacity Act (England & Wales), 2005. Participants were offered a voucher as a thank you for their time and trouble.

### Analysis

We used a reflexive thematic analytic approach (18), then mapped inductive themes to our theoretical framework (15) (Figure 1) to consider how they informed our understanding of diagnostic delay. We exported anonymised, professionally transcribed qualitative interviews to NVivo for initial data management. CCa/MR read data for familiarity, then preliminarily coded five transcripts each. Co-authors second read the transcripts and at an analysis meeting, we explored interpretations, meaning and these initial codes and themes. We used manual coding to engage with the data (16); highlighting relevant text and tagging it with code labels. CCa and EW conducted the analysis supported by MR and other co-authors. We continued coding transcripts, iteratively developing the coding framework (19).

**Figure 1:**
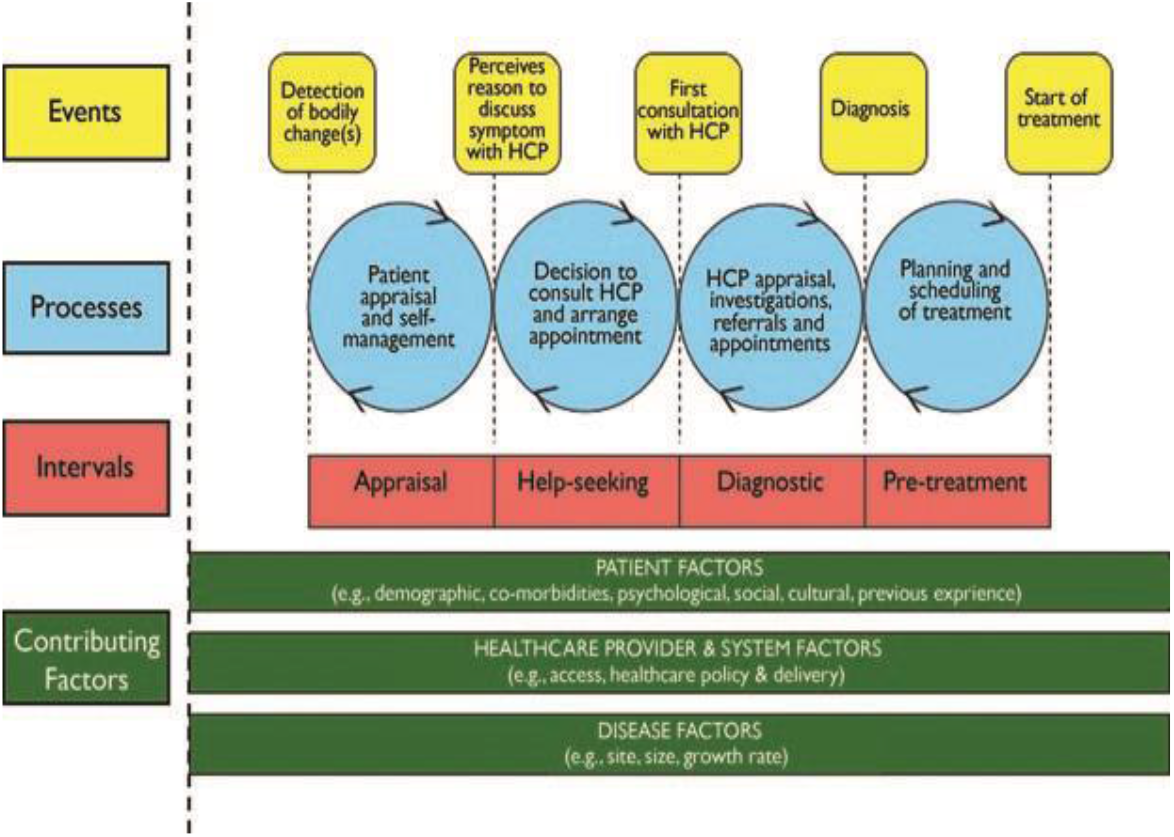
Model of pathways to treatment (15)

At a second analysis meeting we presented partial data utilising 21 anonymised transcripts, to a group including patient and public team members and community stakeholders, welcoming critical debate of emergent themes and new interpretations. The research team used these discussions to refine codes and themes, and led by our study aim, map them to three of the four intervals in our selected theoretical model: appraisal, help-seeking, and diagnosis (15).

## FINDINGS

### Recruitment

61 participants were recruited: 10 individuals living with dementia, 30 family members, 16 health care professionals, 2 interpreters and 3 paid carers. Sociodemographic characteristics, and role characteristics for professionals, are displayed in Table 1.

### Qualitative findings

We identified three themes.

1. **Cultural identity and practices shape responses:** How cultural, family and religious identities and practices shape appraisal and help-seeking.
2. **Becoming like a tourist:** how for some people, developing dementia in an adopted country, engendered a need to reconnect with a home country.
3. **Naming and conceptualising dementia**: exploring how stigma, mistrust and cultural norms might affect timing of a dementia diagnosis

Table three illustrates how our themes informed our understanding of **Appraisal, Help-seeking** and **Diagnostic** intervals as related to our theoretical model.

#### Theme 1: Cultural identity and practices shape responses

This theme represented the centrality of family context to symptom appraisal and help-seeking; family members often held insider and outsider identities, balancing more traditional values and attitudes with support from health care professionals. Gendered role expectations and religious identities shaped how families appraised difficulties and sought and provided support.

For many interviewees, being part of a traditional, often intergenerational household was central to symptom appraisal. Interviewees described how within their local Bangladeshi community, it is considered respectful for a daughter to care for her mother and mother-in-law, even when the older person remained well: preparing food, shopping and sometimes washing and dressing. An interpreter translating for male Bangladeshi family carer, referred to this relinquishing of roles by older women: *“four years ago sister-in-law came and she’s not been cooking since*.*”*

In this context, families sometimes did not realise that their parent had lost skills. A memory clinic nurse described this within multigenerational households:

*“in the Bangladeshi families the custom is that once there is a daughter in law in the household she takes over a lot of the things, so the mother or the person who is being assessed and if it’s a female, it’s almost an expectation that she would lay down her tools and let the young ones get on with things and that can be disabling for people, even though it’s a choice to do it. It means that, it’s difficult for us to know whether the person still can”* (Memory clinic nurse, female, Black African)

Another nurse commented on the challenges of working with families where caring is perceived as a female, and liaising with health care professionals a male role:

*“you ask them, “Well, do you look after mum or dad and stay with them?” And they look at you and say, “Of course I don’t,” it’s like… or, “My wife*.*” And it’s like, “Well, could I speak to them?” And a lot of the time, the answer is yes, but not completely, sometimes “You cannot speak to a female in my family*.*”* (Male consultant liaison nurse, White British)

Family members who were born in Britain to immigrant parents described inhabiting insider/outsider cultural identities, navigating a process of seeking help within a Western healthcare system that is sensitive to their parents or families cultural and ethnic background. This family carer relates her experiences of appraising when to seek help to living in an area with low ethnic diversity:

*“I think because I’m their only child. So, me repeatedly saying something, you could class me as a community because where we live is a predominantly white area. We don’t live in an Asian area. So, if we did, there’s a thing of like other people would have noticed it, then it might have been more of a laughing talking point, “Uncle’s doing this again,” whatever’s doing it. It’s more of a joke*.*”*

(Female family carer, Bangladeshi)

The same family carer explained how people in her community see caring as a duty and did not identify with the label of carer since *‘if you label it, you’re failing’*, but how this may be an obstacle to accessing support.

Changes in religious practice appeared to be events which would trigger a recognition of a cognitive concern and instigate a response. We heard examples of this within the Muslim community in relation to prayer and use of the Quran. It was also evident within the Caribbean community when older members of the church community developed dementia symptoms and were distressed that they were unable to take part as independently as previously. The emotional impact of reduced ability to perform and observe religious roles is evident in the following account from a father, who chose for his daughter to translate:

*“So, he’s saying the fact he forgets, and especially when it comes to prayers, so you have certain times you have to do it, and he forgets whether he’d done it. So he performs it again, then when he’s performing, he makes mistakes as well, so he kind of, he remembers, did I say the specific verse? Then it’s a vicious circle he’s in, so he’s quite muddled in his brain, that’s what he said*”.

(Male diagnosed with dementia, Bangladeshi)

When to pray, the order of prayers and Quranic verses are learnt in childhood, and so to forget was keenly felt as an indicator of cognitive loss, with a fundamental impact on an individual’s role and identity. Religion also affected the appraisal of a situation as the following account illustrates from a health care professional.

*“I’ve seen, and I’ve heard patients talk about their religious beliefs. So that’s kind of also, I would say, a barrier to them reporting symptoms because they think, they believe, that whatever they’re experiencing is related to Allah, kind of directing or giving this to them, and therefore it’s Allah who will heal them, or will help them through this” (Occupational Therapist, female, Pilipino)*

#### Theme 2: ‘You become like a tourist’ immigration experiences and language barriers

This theme encompasses areas linked to familiarity: how experiences and cultural practices which are familiar, can become unfamiliar through dementia and how language barriers can become more disabling when dementia symptoms also challenge communication. It includes examples of people travelling back to a country of origin as part of a family’s response to cognitive concerns. Narratives about movement and travelling were identified across a number of accounts. These were arranged by members of the family members, usually adult children, and all involved travel to a country from which their parents originated, to appraise or seek help for dementia symptoms, with the following quotation illustrating this,

*Yeah, because she wasn’t all that, so I said, “Let me take her to the Caribbean to see if she’s going to be the same. She (Mum) was OK for a while, then she started the same thing/pattern again”*.

*(Female family carer, Black Caribbean)*

Appraising changes would also happen following movement back home and travel.

*“She said that before she can feel that his memory gone but it’s not strong like now. Before that he went to the hospital and they got an ear test done. But since he came back from Vietnam that is where she knew that he lost his memory” (Wife of man with dementia, Vietnamese)*

There were examples of first-generation individuals expressing a desire to return ‘one more time’ to a homeland, when they recognised that their memory was becoming worse. One participant described feeling like a ‘tourist’ due to memory problems after having lived in the UK for fifty years. He felt disorientated in a usually well-known location, which in the interview he compared to his early experience of unfamiliarity after arriving in the UK.

*“I have been living for long here but when this started I become like a stranger, like I just come to this country, now, if I go to the train station, I have to look at the map, you become a tourist”*

(Man with dementia, Caribbean background)

A manager of a centre described how important trust was to older people from the Caribbean community, suggesting that the yearning for home country might also be met through availability of specific cultural social resources.

*“They trust us. They trust us, and they could see how we are with their family members. And as I said, we go above and beyond to help and engage with them. And I think they probably need more out there. More, yes*.*” (Manager, female Black Caribbean)*

Language differences created challenges for people with dementia their families and health care professionals to ensure that information is accessible, and concerns and needs are voiced, heard, and understood. Translation may not convey meaning, and professionals and interpreters described challenges in balancing voices of relatives and clients. A GP described the challenges of translating not only words but meaning:

*“the translation of the word dementia, when I say to the interpreter, can you tell this person they have dementia. I’m not really sure how, I am describing it and dementia means that. I am getting those bits translated but the word in itself, I’m not convinced that there is a word, I’m not convinced that there is a selective word that completely is a direct translation”*

*(Consultant psychiatrist, Male, White British)*

There was a sense in several accounts of a “double jeopardy” of communication barriers for people with dementia accessing healthcare via an interpreter, especially where accompanying relatives are able to engage directly with professionals in English. A family carer (female, Bangladeshi) stated that both her parents had symptoms consistent with dementia but only her father had a diagnosis, and she attributed this to his ability to speak English, while her mother did not. She felt that there was difference in how they were heard, and felt that the translator was not conveying everything reliably from her mother to the doctor in the memory clinic:

*“I mean, my personal view is, if you can speak English well, you have one service. If you can’t, you have a totally different one”. (Daughter and carer, Bangladeshi)*

In the example below a son felt his Mum was repeating things or ‘*telling too many stuff’* and therefore intervened through translating parts of the exchange.

*“No, she doesn’t go out, so basically even with an interpreter sometimes we still need to help her when she is seeing the doctor, she doesn’t remember, or she keep repeating. Or she probably telling too many stuff at the same time. We have to assist her even with the interpreter, she is fully dependent on us”* (Male family carer, Bangladeshi background)

Health care professionals described challenges accessing professional interpreters, leading to use of telephone interpreting services. One GP felt this increased the risk of misunderstandings, as non-verbal cultural nuances were lost.

#### Theme 3: Naming and conceptualising dementia

In this theme, we discuss how the term dementia was stigmatised and there were cultural nuances to how it was understood. This theme held relevance across the three stages of our conceptual model – appraisal, help-seeking and diagnostic intervals.

Cognitive symptoms were the primary trigger for help seeking for the majority of the people living with dementia and family carers that we interviewed; however, other symptoms were present and were a trigger for help-seeking for some. Health care professionals described how consultations with reoccurring physical concerns such as mobility pains and arthritis could mask dementia symptoms, extending the diagnostic interval. Several GPs interviewed described this, including the quotation below:

*“they do come with vaguer symptoms sometimes, like it’s often – yes, actually, I guess when they’re coming indirectly, it’ll be repeated appointments for the same thing, or sort of a preoccupation with an arthritic knee, something just coming again and again, and eventually, and you think, “Well, something’s not right here*.*” I had one patient like that, and it did take me a while for the penny to drop”* (GP, female, described herself as mixed background)

Families also often sought help in this way, attributing behavioural or cognitive changes to physical illness, or sought help for their relative’s physical health problems, because they did not feel able to name cognitive symptoms, as in this quote:

*“so they won’t have been told it’s because of a memory problem that they’re coming to see the doctor. They’ll have been told you’re coming about the cough that you’ve had, or whatever it is, So then they want to tell you about that, and that’s why it’s like shh, shh, no don’t worry they’re just trying to tell you about the cough they’ve got. And it’s – so it will be brought under like under sort of false pretences to come and see me*.*” (GP, female, white British)*

A neurologist attributed initial presentations often including physical symptoms with cognitive concerns to a reluctance to address mental health issues, including cognitive concerns, which as the quote below illustrates, could extend the appraisal and help-seeking intervals:

*“from personal experience as an Asian person with an Asian family, I know that there is a huge emphasis on physical health, and it’s very, very difficult, I think, to talk to people about mental health, and even the idea that it could have aspects impact on physical health. So I don’t think it’s that the patients biologically are obviously presenting differently, I think it’s that it’s what they are noticing and wanting to seek help for is different, and similarly, what their family are noticing and wanting to seek help for is different. “(*Consultant neurologist, male, Sri Lankan)

Many accounts described the terms dementia and forgetfulness being used interchangeably, with an implication that dementia is a normal part of ageing, but this was culturally specific. These included an account from a daughter caring for her father, from a Bangladeshi background, who commented about dementia: “*essentially, it’s forgetting”*. This Vietnamese interpreter described how cultural norms influenced how she translated during diagnostic appointments:

*“the translation for dementia in Vietnamese is very … basically, you’re forgetful, they don’t use that word ‘dementia” (Female interpreter in memory clinic, Vietnamese)*

Several interviewees described milder forms of dementia as “forgetting”, while reserving the term dementia for severe dementia. A GP observed that for her patients, the term dementia was reserved for severe cognitive impairment, with an implication that mild or even moderate dementia was not accepted as such:

*“I think because I am from Trinidad it is fair for me to talk about it, so the patients I would be able to say to them, yes I get it, I get what you see as dementia. I know what dementia means when you are in the Caribbean, I don’t know how to phrase this in a correct way, but really completely lost all their cognitive functions, is what they see dementia as”*. (GP, female)

These appraisals potentially extended the help seeking interval. Many interviewees identified a discomfort with the word dementia with one female Bangladeshi family carer referring to both dementia and mental illnesses such as depression as being ‘taboo’ in their culture, particularly for the older generation: “*Mental health is taboo. You just don’t talk about it at all”*

This seemed to reflect a sense of shame around cognitive decline, poor mental health, and dementia, which although rarely voiced by patients or family carers, was referred to by several healthcare professionals, including in the extract below:

*“different groups might feel less trust for health professionals and obviously going to the GP about your memory is really exposing. And there is lots of fears of stigma and not being taken seriously and even having your rights removed and what’s going to happen if I open up about this for myself”*

*(GP, female, White British)*

A family carer also linked reluctance to seek help to trust, commenting: :

‘if *you don’t really “recognise/understand dementia conceptually and there is a stigma in it, then why would you seek help”* (Family carer, female, Bangladeshi)

Another also linked an extended help-seeking interval to low expectations of the care they would receive and an absence of a ‘mental model’ of help-seeking:

*“And there isn’t a preventable attitude in the countries that don’t have a free healthcare system. If you’ve come from somewhere that doesn’t have social services, doesn’t have a free healthcare system, you are not going to get intervention because you have to pay for it. So, you don’t have a natural mental model to go,”* (Female family carer, Bangladeshi)

## DISCUSSION

We identified three themes responding to our aim, to explore the pathways determining timing of dementia diagnoses in an ethnically diverse population. They explore how cultural identity and practices shape responses to dementia symptoms, with second generation family members often balancing Western care systems with traditional values and attitudes, including gendered expectations around household roles, and the importance for some communities of religious practice in evaluating functioning in many communities. The onset of dementia held particular challenges for first generation immigrants, for whom it sometimes engendering a need to reconnect with a home country, while for those for whom English was not a first language, linguistic and dementia-related communication barriers could cause a “double jeopardy” in terms of communicating with health and care providers. Interpreters described the challenges in translating meanings as well as words, and balancing relatives’ and clients’ voices. Discourses around stigma were evident in accounts, and this and feelings of mistrust in services proposed to explain the common observation that initial help seeking was often for physical symptoms.

Our themes were all relevant to the appraisal interval of the Model of Pathways to Treatment (16) (Table 3). This posits that people decide to consult a healthcare professional about a symptom when they believe something is wrong or serious, or that the problem is interfering with their functioning and beyond their ability to cope. Our findings echo previous reports that in South Asian communities, ceding household duties to a daughter-in-law, meant loss of such skills was not considered problematic (11). By contrast, decline in religious practice often indicated to families that help was needed. This was reported by Muslim and Christian participants and is not to our knowledge, previously discussed in the literature. Notions of karma have been cited to explain how Hindu participants understood and responded, with acceptance to dementia (9,10).

Scott et al (16) discuss how expectations of how services might help determine ‘Appraisal’ and ‘help seeking’ intervals. Beliefs that seeking help will reassure, help symptoms, or improve prognosis promote early help seeking. Negative expectations of unwanted treatment, investigations, embarrassment, or disruption to self-esteem, self-identity or sense of independence, delay it. For many of our interviewees, stigma seemed to underlie delayed help seeking, or presentations with physical rather than cognitive symptoms, due to embarrassment. Among minority ethnic carers in England, stigma and mistrust of services, and beliefs that physical illnesses, feigning or life changes were causing dementia symptoms are previously cited as reasons for delayed presentation (11). Unlike in this previous study, our findings suggested presentations with physical symptoms were strategies to raise cognitive concerns despite stigma, rather than due to a lack of attribution to dementia.

We identified the insider/outsider roles held by family carers as enabling help-seeking and obtaining a diagnosis. In previous work, family carers from Bangladeshi and Indian ethnic backgrounds in England have described wanting to be supported by services, but encountering unacceptable care models, and tensions between expectations of first generation parents and their other, work and family roles (21). Previous research has not distinguished between the experiences of people living in areas of high versus low ethnic diversity: these may differ, as described by a family carer participant who felt her situation would be different if supported by a larger local community. According to Social Cognitive Theory, social contacts influence when and whether an individual decides to seek care (16). This aligns with findings regarding adverse mental health outcomes, with those in areas with low ethnic diversity experiencing exclusion from local networks, geographically dispersed culturally specific services, and more damaging effects from everyday racism (22). Perhaps this need to consult trusted social contacts explained why some people developing dementia and their families wanted to return to a home country to inform how they appraised symptoms. This concept, and notions of feeling “like a tourist” has not to our knowledge been described previously in the published dementia literature, though Roche in his thesis describes a similar theme denoting a state of precarity in the sense of belonging and social position Black African and Caribbean people with dementia experience in UK (28).

Previous authors have described a need for ethnic and language matching between interviewers and participants in research in minority ethnic communities to elicit trust and shared understanding (24), however the interviewers in this study had different ethnic backgrounds (Black Caribbean and White British) to the interviewees. Qualitative studies of this nature risk unintentionally “othering” minoritized ethnic groups in a way that is unhelpful and inaccurate. Our intention was not to compare pathways to help seeking in minoritized ethnic communities to those of other groups, but to understand why people from minoritized ethnic groups receive dementia diagnoses later, and how services could be adapted to account for different styles of appraisal and help-seeking to reduce this inequality. For example, the belief that dementia is part of normal ageing may be no more common in minoritized ethnic than majority ethnic populations (23). Normalising bodily changes saves cognitive effort and allows functioning to continue (25); as people grow older, they increasingly attribute sensations to ageing rather than illness (26). Individuals across all cultures who are embarrassed, frightened or ashamed by symptoms will commonly delay seeking help (27). But our finding that fear of stigma, and a sense of alienation from services in first generation immigrants might underlie these processes for some minoritized people can inform interventions to reduce inequalities of experience.

This is to our knowledge the first qualitative study exploring how people from ethnically minoritized communities seek help for dementia symptoms to include perspectives of people with undiagnosed dementia and interpreters. The diagnostic interval was affected for many interviewees by the complexities of translating and interpretation. Previous studies, which interviewed health care professionals but not interpreters, described difficulties assessing dementia due to language barriers, including interpreters changing sentences or meanings to facilitate the translation and thus introducing errors (6).

Our findings support calls for culturally sensitive health promotion interventions, more minority ethnic staff in services, staff to be culturally competent and use of interpreters with some dementia training (23). They suggest some common practical dilemmas for families that might inform interventions. The need for strategies to raise cognitive impairment sensitively in different cultures has been previously described in Norway, where driving-licence status is commonly used to address cognitive symptoms among the majority population, but less helpful for older immigrants who less often drive (6). Perhaps for some communities, religious activities, rather than household tasks, for which families compensate for functional decline with collectivistic approach to support and care (6), are a helpful focus when exploring functioning. Our findings suggest that clinicians and interpreters planning in advance how to convey diagnosis and key information could be factored into clinic timings, acknowledging that providing high quality, respectful dementia care through an interpreter takes longer.

Our findings suggest that clinicians aim to plan how best to use interpreters and avoid phone translation where possible. Ideally clinicians and interpreters should plan in advance how to convey diagnosis and key information could be factored into clinic timings. Clinicians should consider the cultural variations of symptom attribution which includes a personal and social construction of meaning of dementia and how this affects the clinical presentation of cognitive concerns. This includes the physicality of cognitive health concerns and the normalisation through role and beliefs.

## Data Availability

All data produced in the present study are available upon reasonable request to the authors

## Authors Contribution

CM is the chief investigator of the study and secured the funding for the study; CCa and CCo drafted the manuscript. JB managed, and CCa and MR carried out recruitment and study procedures. All authors contributed intellectual content to the study design, paper and reviewed the final version prior to submission.

